# Quantitative and radiological assessment of post-cardiac arrest comatose patients with diffusion-weighted magnetic resonance imaging

**DOI:** 10.1101/2023.11.29.23299196

**Authors:** Sam Van Roy, Liangge Hsu, Joseph Ho, Benjamin Scirica, David Fischer, Samuel Snider, Jong Woo Lee

**Affiliations:** Department of Neurology, Division of EEG and Epilepsy, Brigham and Women’s Hospital, Boston, MA; Department of Radiology, Division of Neuroradiology, Brigham and Women’s Hospital, Boston, MA; Department of Medicine, Division of Cardiology, Brigham and Women’s Hospital, Boston, MA; Department of Neurology, Division of Neurocritical Care, Philadelphia, PA; Department of Neurology, Division of Neurocritical Care, Brigham and Women’s Hospital, Boston, MA

**Keywords:** Cardiac Arrest, Anoxic Brain Injury, Coma, Neurological Recovery, Diffusion Weighted Imaging, Radiological Assessment

## Abstract

**Background:** Although MR imaging, particularly diffusion weighted imaging, has increasingly been utilized as part of a multimodal approach to prognostication in patients comatose after cardiac arrest, the performance of quantitative analysis of apparent diffusion coefficient (ADC) maps, as compared to standard radiologist impression, has not been well characterized. This retrospective study evaluated quantitative ADC analysis to the identification of anoxic brain injury by diffusion abnormalities on standard clinical MRI reports.

**Methods:** The cohort included 204 previously described comatose patient post-cardiac arrest. Clinical outcome was assessed by 1) 3-6 month cerebral performance category (CPC); 2) Coma recovery to following commands. Radiological evaluation was obtained from clinical reports and characterized as diffuse, cortex only, deep gray matter structures only, or no injury. Quantitative analysis of ADC maps were obtained in specific regions of interest (ROI), whole cortex, and whole brain. A subgroup analysis of 172 was performed after eliminating images with artifacts and pre-existing lesions.

**Results:** Radiological assessment outperformed quantitative assessment over all evaluated regions (AUC 0.80 for radiological interpretation, 0.70 for occipital region, the best-performing ROI, p=0.11); agreement was substantial for all regions. Radiological assessment still outperformed quantitative analysis in the subgroup analysis, though by smaller margins, and with substantial to near-perfect agreement. When assessing for coma recovery only, the difference was no longer significant (AUC 0.83 vs 0.81, p=0.70).

**Discussion:** Although quantitative analysis eliminates interrater differences in the interpretation of abnormal diffusion imaging and avoids bias from other prediction modalities, clinical radiologist interpretation has a higher predictive value for outcome. This difference appears to be driven by poor scan quality, foreign body artifacts, and pre-existing stroke and white matter disease. Quantitative analysis is comparable to clinical interpretation after eliminating such scans. Further research is required into improving quantitative imaging techniques to account for such variability.

## Introduction

Neuroprognostication after cardiac arrest remains a challenging but critical aspect in the management of comatose patients after cardiac arrest, as most mortality in-hospital is due to withdrawal of life-sustaining treatment due to poor neurological prognosis [1]. Since its clinical introduction as a tool for early stroke detection in the early 1990s, diffusion-weighted imaging (DWI) has evolved to complement other prognostic markers, including EEG, SSEP, and neuron-specific enolase [2–6].

DWI can be utilized to predict neurological outcome with high accuracy, but there is limited generalizability in the current methods due to differences in a variety of factors, including lack of a standard interpretation [7]. Clinical radiology interpretation remain the basis for DWI interpretation, but may be limited by interrater variability, lack of objective cut-off measures for both severity of injury, and knowledge of other clinically relevant prognostic information, potentially resulting in a self-fulfilling prophecy bias. Quantitative measures utilizing calculated apparent diffusion coefficient (ADC) maps have been utilized to minimize these limitations [8–13].

Previous studies revealed that clinical radiologist interpretation of the MRI outperformed quantitative ADC map analysis in outcome prognostication [9,14]. This study aims to compare quantitative ADC map analysis with the clinical radiology interpretation of ADC diffusion restriction to assess the degree of agreement and correlation with clinical and neurological outcome, and to evaluate potential sources of discrepant findings.

## Methods

### IRB Approval

This study was approved by the Mass General Brigham institutional review board under protocols 2014P001623 and 2021P000633. The requirement for participant consent was waived for the study.

### Description of Participants

A retrospective selection of 204 patients was done from a Brigham and Women’s Hospital registry of in-hospital and out-of-hospital cardiac arrest patients between 2009 and 2020 from a previously described dataset [8]. In brief, patients were over 18 years old, unresponsive upon first assessment after the return of spontaneous circulation, and had MRI performed within 14 days post-cardiac arrest.

Basic demographic information, the time between cardiac arrest and acquisition of DWI, radiology reports, cerebral performance scale (CPC) scores obtained 3-6 months post-cardiac arrest, coma recovery to following commands, the use of Targeted Temperature Management (TTM), were extracted from the hospital’s electronic medical record system (Table 1).

**Table 1.** Cohort characteristics and demographics. CI, confidence interval; TTM, targeted temperature management. (Taken from Snider et al., 2022)

|  |  |
| --- | --- |
|  | Patients with<br>cardiac arrest (n =<br>204) |
| Mean age (95%<br>CI), y | 55 (53, 58) |
| Male sex, n (%) | 131 (64) |
| White ethnicity, n<br>(%) | 130 (64) |
| Received TTM, n<br>(%) | 176 (86) |
| Followed<br>commands before<br>discharge, n (%) | 80 (39) |
| Died before<br>discharge, n (%) | 116 (58) |
| Mean days between<br>arrest and MRI<br>(95% CI) | 5.0 (4.6, 5.3) |

### MRI Processing

MRI acquisition and post-processing was performed as previously described.[8] In brief, the initial post-cardiac arrest diffusion and T1 sequences were used for analysis. All acquired diffusion sequences included b = 0, 500, 1,000 s/mm^2^, 1.5 x 1.5 x 6 mm voxel size, and diffusion encoding along the principal axes. T1-weighted sequences included sagittal and axial pre-contrast volumes (voxel size ∼ 0.5 x 0.5 x 5 mm) or a magnetization-prepared rapid gradient echo volume (voxel size 1 x 1 x 1 mm).

Quantitative analysis of the ADC maps was performed on diffusion-weighted sequences. ADC maps were registered into standard Montreal Neurological Institute 152 space (MNI space) by sequentially performing brain extraction (FSL/bet2), followed by linear, and nonlinear diffeomorphic registration (ANTs:antsRegistrationSyNQuick). When a T1-weighted isotropic magnetization-prepared rapid gradient echo sequence was unattainable or not produced, the clinical sagittal and axial T1 volumes were used to construct a higher resolution volume using NiftyMIC (Ebner et al., 2020). Every registration was also inspected manually via slicesdir (FSL 6.0.1), and two MRIs were excluded due to the registrations of gross inaccuracy. ADC maps underwent a rigid body registration to the T1 weighted image, then the above transformations were then applied to register them into the Montreal Neurological Institute space for comparison.

The ADC maps excluded values outside the range of 200 to 1,200 x 10^−6^ mm^2^/s to mitigate artifact and cerebrospinal fluid signal contribution. The threshold used was based on previous works done in the field [9,11]. Mean ADC values of specific regions of interest (ROIs) were calculated for each patient. This was performed by transforming a priori pre-defined ROIs in MNI space onto each patient’s ADC map. ROIs for gray matter were the bilateral frontal, temporal, insular, parietal, and occipital lobes, and the cerebellum based on the MNI Structural Atlas [15]. Gray matter ROIs also included the bilateral basal ganglia (including the caudate, putamen, and pallidum), bilateral thalamic, and brainstem based on the Harvard-Oxford Subcortical Atlas [16]. White matter ROIs included the bilateral and binarized tracts and the corpus callosum based on the Johns Hopkins University White Matter Atlas [17] and the Jülich Atlas [18] respectively.

Due to known factors confounding DWI scans, a subgroup analysis was performed after eliminating scans that were determined to have had either poor quality or artifactual scans or pre-existing abnormal ADC values by the study investigators (SV, JWL). These included image artifacts (shunts, metallic objects, others), prior strokes (as evidenced on imaging prior to cardiac arrest or encephalomalacia on CT/MRI), and severe white matter disease (diffuse FLAIR-bright white matter lesions). To minimize bias in selection, the investigators were blinded to outcome measures when selecting the subgroup.

Clinical MRI interpretations were obtained from the patient’s medical records. Radiology reports were categorized utilizing the following grading system based on assessment of diffusion restriction: grade of 1: diffuse anoxic injury (both cortical and subcortical structures); grade 2: cortical anoxic injury only; grade 3: subcortical structures: thalamus, basal ganglia, cerebellum, injury without cortical involvement; grade 4: no anoxic injury. No additional assessments were performed outside of the clinical radiological reports. This grading system was chosen to reflect severity of injury, similar to previously published criteria [14,19] and adapted to allow extraction of elements from clinical neuroradiological interpretation.

### Evaluation and Classification of Outcome

Separate outcome analyses were performed with 2 different classifications of good vs poor neurologic outcomes. Long-term neurological evaluation scores measured 3-6 months post-cardiac arrest were obtained utilizing the cerebral performance category (CPC) scale. Shorter-term coma recovery was assessed by the ability of the patient to follow commands at any time prior to hospital discharge. Outcome group 1 defined good neurologic outcome as a CPC of 1-2, indicating independent function. Outcome group 2 defined a good outcome as a patient who followed commands upon assessment; this was performed to account for patients who may have good neurological recovery but subsequently died due to other non-neurological clinical factors.

### Statistical analysis

To assess the degree of agreement and correlation with neurologic clinical outcome of the quantitative dataset and the clinical radiological interpretations, separate logistic regression models for each outcome group and region of interest were constructed. Quantitative models were performed at an ADC range to 200 – 1200 x 10^−6^ mm^2^/s. Receiver operating characteristic (ROC) curves were generated from the R package pROC [20] and ROCR [21]. Sensitivity, specificity, positive predictive value (PPV), and negative predictive value (NPV) were calculated based on the optimal threshold determined from the ROC analysis.

Delong’s test was used to compare the ROC curves of each model. Cohen’s kappa value was used to measure agreement between models at the optimal threshold in both parts of our analysis. Agreement was considered none/slight (Kappa 0.01-0.20), fair (0.21-0.40), moderate (0.41-0.60), substantial (0.61-0.80), or almost perfect (0.81-1.00) [22] All statistical tests were considered significant at an α < 0.05 level. A subgroup analysis was similarly performed excluding patients with the previously described confounded variables.

## Results

A total of 206 patients were included in the study. Patient characteristics are shown in Table 1. The mean time post-cardiac arrest and MRI was 5 days. A total of 39% of patients followed commands and 58% of patients died before discharge.

Analysis of the comprehensive dataset is shown in Table 2. In Outcome Group 1 AUC was 0.80 for radiological interpretation, 0.70 for the best-performing region (occipital lobe, p = 0.011 as compared to radiological interpretation), 0.66 for whole cortex (p = 0.0015), and 0.63 for whole brain (p < 0.0005) (Figure 1). For Outcome Group 2 AUC was 0.81 for radiological interpretation, 0.77 for the best-performing region (occipital lobe, p = 0.17), 0.72 for whole cortex (p = 0.020), and 0.70 for whole brain (p = 0.0050). Cohen’s kappa values were substantial for almost all Outcome groups and regions (Table 2).

**Table 2.** Comprehensive dataset results. Area under the curve (AUC), sensitivity, specificity, optimal threshold, Cohen’s kappa value compared to the radiological interpretations, positive predictive value (PPV), and negative predictive value (NPV) are reported for the comprehensive dataset.

| Region/Model | Outcome Group 1 |  |  |  |  |  |  | Outcome Group 2 |  |  |  |  |  |  |
| --- | --- | --- | --- | --- | --- | --- | --- | --- | --- | --- | --- | --- | --- | --- |
|  | AUC | Sensitivity | Specificity | Optimal Threshold | Cohens Kappa | PPV | NPV | AUC | Sensitivity | Specificity | Optimal Threshold | Cohens Kappa | PPV | NPV |
| Radiological Interpretation | 0.80 | 0.89 | 0.66 | 4.00 | NA | 0.92 | 0.51 | 0.81 | 0.78 | 0.81 | 4.00 | NA | 0.86 | 0.72 |
| Basal Ganglia | 0.60 | 0.64 | 0.47 | 771.50 | 0.63 | 0.77 | 0.30 | 0.65 | 0.73 | 0.55 | 769.66 | 0.65 | 0.77 | 0.50 |
| Thalamus | 0.52 | 0.68 | 0.38 | 798.65 | 0.54 | 0.78 | 0.28 | 0.55 | 0.58 | 0.53 | 815.47 | 0.53 | 0.67 | 0.43 |
| Frontal Lobes | 0.65 | 0.68 | 0.49 | 827.29 | 0.63 | 0.84 | 0.35 | 0.69 | 0.63 | 0.67 | 841.69 | 0.62 | 0.75 | 0.54 |
| Occipital Lobes | 0.70 | 0.57 | 0.67 | 865.31 | 0.69 | 0.81 | 0.39 | 0.77 | 0.81 | 0.65 | 834.39 | 0.72 | 0.85 | 0.59 |
| Parietal Lobes | 0.66 | 0.75 | 0.49 | 832.02 | 0.69 | 0.85 | 0.36 | 0.73 | 0.80 | 0.60 | 830.88 | 0.69 | 0.83 | 0.55 |
| Temporal Lobes | 0.61 | 0.71 | 0.48 | 848.76 | 0.64 | 0.82 | 0.33 | 0.66 | 0.72 | 0.54 | 846.31 | 0.64 | 0.76 | 0.50 |
| Whole Cortex | 0.66 | 0.68 | 0.57 | 850.00 | 0.68 | 0.83 | 0.37 | 0.72 | 0.78 | 0.60 | 836.25 | 0.68 | 0.82 | 0.55 |
| Whole Brain | 0.63 | 0.64 | 0.55 | 828.61 | 0.66 | 0.81 | 0.34 | 0.70 | 0.75 | 0.60 | 820.02 | 0.66 | 0.79 | 0.54 |

**Figure 1.**
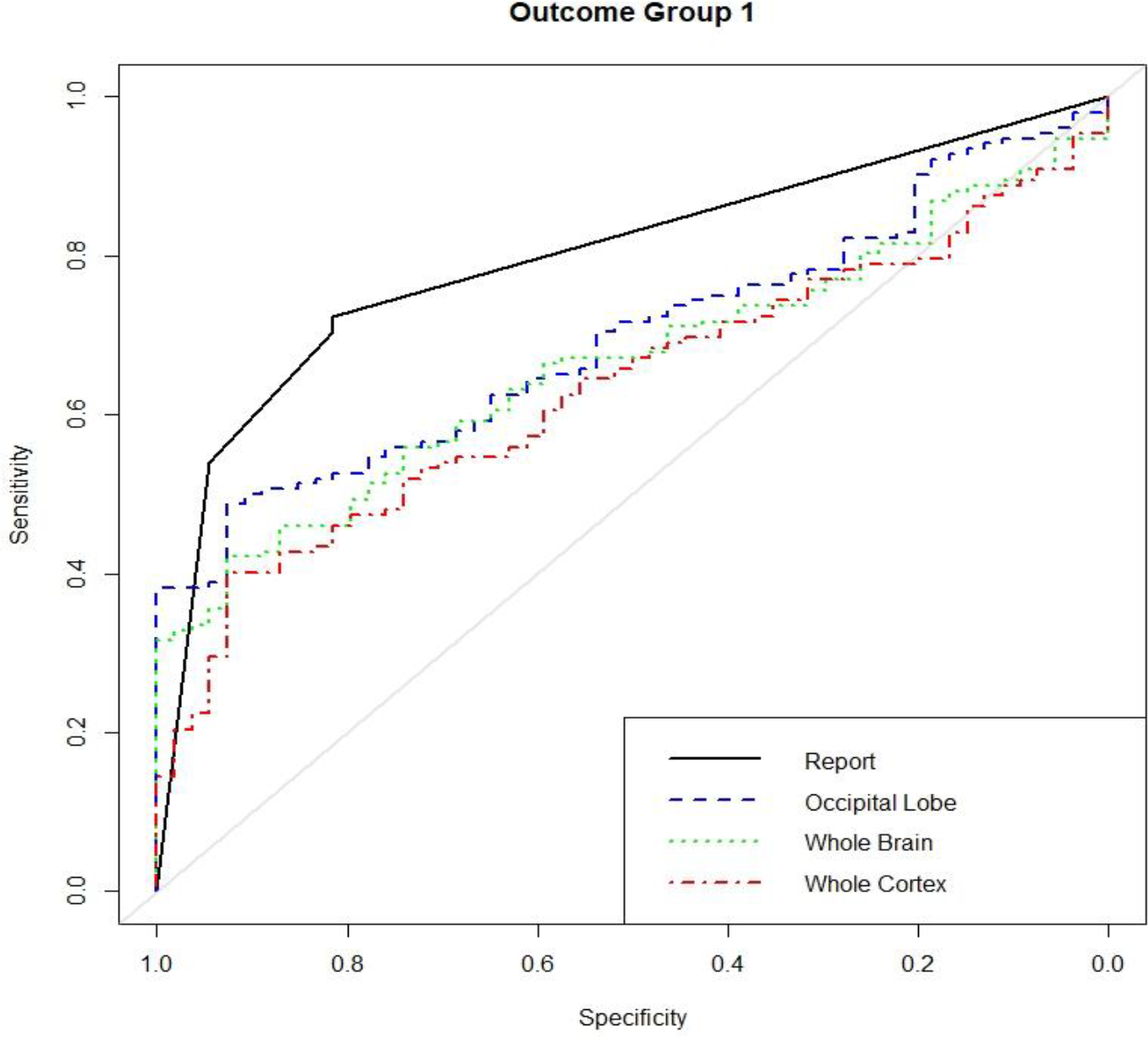
Comprehensive dataset Outcome Group 1 receiver operating characteristic curves. Radiological report (black) AUC was significantly greater than the occipital lobe ADC (blue), whole brain ADC (green), and whole cortex ADC curve (red) (p < 0.05).

In the subgroup analysis, a total of 14 patients were eliminated due scanner artifacts (most comm only metal induced), 10 for prominent white matter disease, 7 for prior strokes, 2 for excessive patient movements, and 1 for excessive calcifications (34 total exclusions).

Analysis of the subgroup dataset (172 patients) is shown in Table 3. In Outcome Group 1 AUC was 0.84 for radiological interpretation, 0.76 for the best-performing region (occipital lobe, p = 0.031 as compared to radiological interpretation), 0.72 for whole cortex (p = 0.0051), and 0.69 for whole brain (p = 0.00089). For Outcome Group 2 AUC was 0.83 for radiological interpretation, 0.81 for the best-performing region (occipital lobe, p = 0.70), 0.76 for whole cortex (p = 0.10), and 0.73 for whole brain (p = 0.030) (Figure 2). Cohen’s kappa values were substantial to near perfect for all Outcome groups and regions (Table 3).

**Table 3.** Subgroup dataset results. Area under the curve (AUC), sensitivity, specificity, optimal threshold, Cohen’s kappa value compared to the radiological interpretations, positive predictive value (PPV), and negative predictive value (NPV) are reported for the subgroup dataset.

|  | Outcome Group 1 |  |  |  |  |  |  | Outcome Group 2 |  |  |  |  |  |  |
| --- | --- | --- | --- | --- | --- | --- | --- | --- | --- | --- | --- | --- | --- | --- |
| Region/Model | AUC | Sensitivity | Specificity | Optimal Threshold | Cohens Kappa | PPV | NPV | AUC | Sensitivity | Specificity | Optimal Threshold | Cohens Kappa | PPV | NPV |
| Radiological Interpretation | 0.84 | 0.88 | 0.69 | 4.00 | NA | 0.92 | 0.60 | 0.83 | 0.77 | 0.84 | 4.00 | NA | 0.86 | 0.75 |
| Basal Ganglia | 0.65 | 0.62 | 0.53 | 771.50 | 0.64 | 0.82 | 0.39 | 0.68 | 0.71 | 0.59 | 765.18 | 0.75 | 0.77 | 0.52 |
| Thalamus | 0.50 | 0.65 | 0.40 | 798.65 | 0.75 | 0.76 | 0.31 | 0.55 | 0.68 | 0.47 | 802.14 | 0.81 | 0.70 | 0.45 |
| Frontal Lobes | 0.69 | 0.73 | 0.50 | 822.64 | 0.84 | 0.85 | 0.40 | 0.71 | 0.73 | 0.58 | 822.64 | 0.80 | 0.78 | 0.52 |
| Occipital Lobes | 0.76 | 0.65 | 0.65 | 851.84 | 0.81 | 0.84 | 0.47 | 0.81 | 0.79 | 0.72 | 834.39 | 0.81 | 0.84 | 0.63 |
| Parietal Lobes | 0.71 | 0.73 | 0.55 | 838.63 | 0.81 | 0.85 | 0.42 | 0.76 | 0.86 | 0.60 | 813.74 | 0.79 | 0.88 | 0.58 |
| Temporal Lobes | 0.66 | 0.69 | 0.51 | 848.76 | 0.78 | 0.82 | 0.38 | 0.68 | 0.76 | 0.55 | 840.03 | 0.84 | 0.78 | 0.51 |
| Whole Cortex | 0.72 | 0.65 | 0.62 | 850.00 | 0.84 | 0.83 | 0.44 | 0.76 | 0.76 | 0.65 | 836.25 | 0.69 | 0.81 | 0.57 |
| Whole Brain | 0.69 | 0.69 | 0.55 | 820.02 | 0.84 | 0.84 | 0.42 | 0.73 | 0.71 | 0.65 | 820.02 | 0.84 | 0.78 | 0.56 |

**Figure 2.**
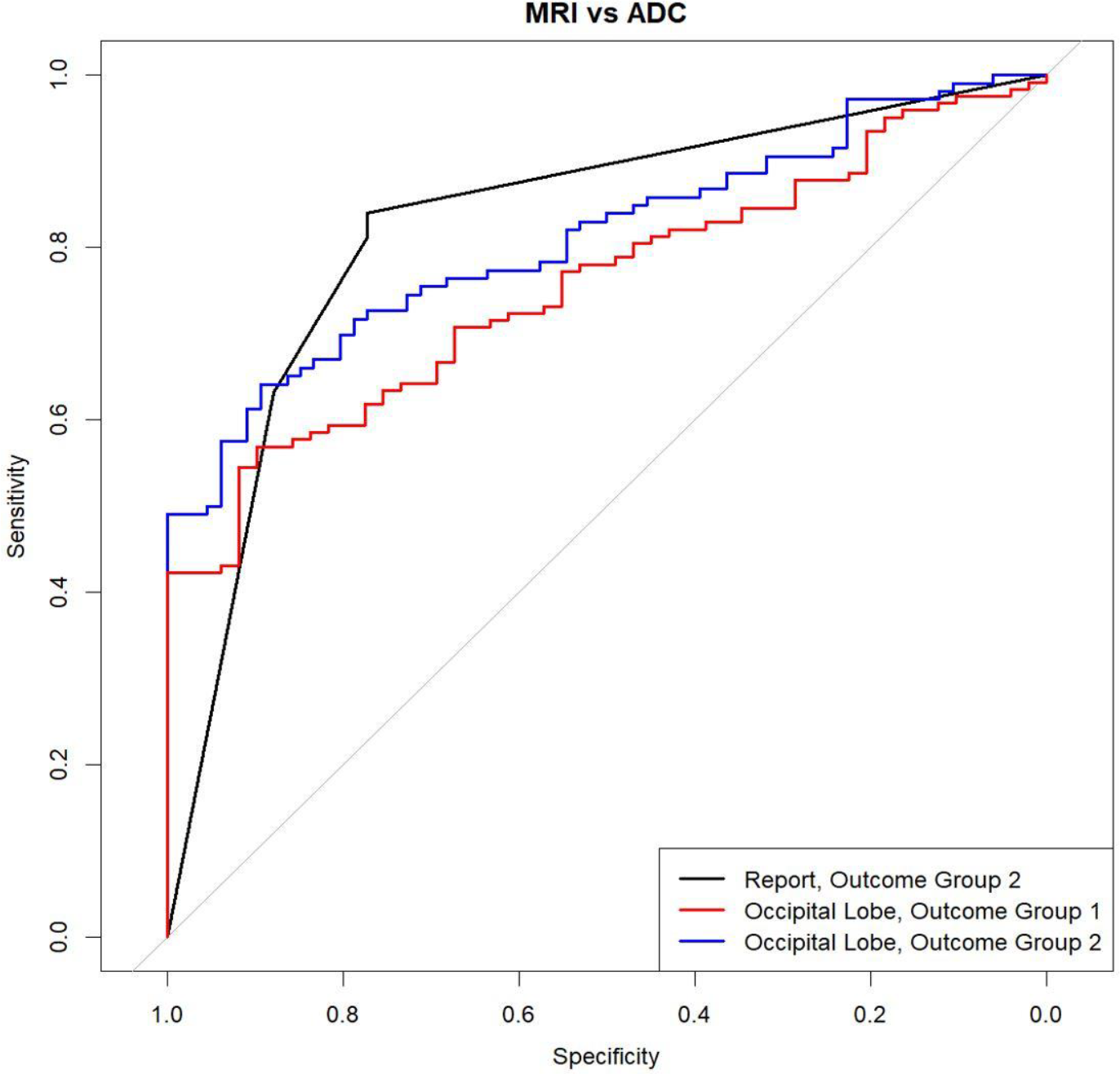
Subgroup dataset receiver operating characteristic curves for radiological report (Outcome Group 1) and occipital lobe ADC data (Outcome groups 1, 2). Radiological report (black) AUC was significantly greater than Outcome Group 1 Occipital lobe AUC (red) (p < 0.05), however was not significantly greater than Outcome Group 2 (blue)

## Discussion

When assessing long-term outcome in patients comatose after cardiac arrest, clinical interpretation of diffusion imaging by a radiologist significantly outperformed quantitative analysis of ADC values of whole brain, whole cortex, or best performing (occipital lobe) ROI (Figure 1). Agreement with radiological interpretation was in the low substantial range (Cohen’s Kappa 0.54-0.69). When assessing for coma recovery to follow commands, there were improvements in the quantitative ADC analysis in all ROIs, radiological interpretation significantly outperformed quantitative analysis in all ROIs except the occipital region. The subgroup analysis performed after removing images with pre-existing lesions and scanner artifacts resulted in small improvement in the clinical and larger improvement in the quantitative analysis; when assessed for coma recovery, the differences were no longer significant, though quantitatively still favoring radiological interpretation (Figure 2). There was also greater agreement between radiological and quantitative analysis (Cohen’s Kappa 0.69-0.84 for coma recovery).

Radiological interpretation may have greater association with outcome as compared to quantitative analysis for several reasons: 1) Radiological interpretation is likely superior to quantitative analysis when there are pre-existing findings and scanner artifacts; in the subgroup analysis which tried to account for this scenario, there may still be artifacts that degraded quantitative performance; 2) Radiologists have access to other sequences whereas quantitative models only used ADC maps; although we limited our findings to radiology reports of diffusion restricting abnormalities, other sequences may have driven attention to certain regions or more subtle findings; 3) Radiological interpretation may be subject to anchoring bias; radiologists are typically aware of clinical findings, CT scans, and other information that may have influenced their interpretation; 4) Radiological interpretation is more susceptible to the self-fulfilling prophecy; clinical decision-making was likely influenced by radiological interpretation, resulting in withdrawal of life-sustaining treatment.

The study has several strengths. The cohort is large, utilizing a previously validated image analysis techniques. ADC values and processing were performed in a uniform manner using standard image processing techniques. To capture the world variability and bias most closely, radiological reports, rather than a review by a blinded radiologist was utilized. Both long-term clinical outcome based on CPC scores and coma recovery based on following commands revealed consistent findings. As patients may expire due to systemic disease unrelated to neurological recovery, the increased correlation utilizing the outcome of coma recovery was expected and in the evaluation of imaging techniques, is a more valid outcome than CPC score. A further analysis binarizing outcome to CPC 1-3 vs 4-5 was performed as it had previous been utilized to asses survival [23]; results were nearly identical to coma recovery, as most patients with CPC3 were able to follow commands (result not shown).

There are several limitations in this study. Grading of radiological interpretations were more elemental than previously described [24]. This 4-point grading system was chosen to allow for data elements to be comprehensively collected from clinical reports. The scale has content validity and has excellent performance on the ROC curve. The subgroup analysis was performed because severe white matter disease, chronic stroke, and calcifications result in elevation of ADC values unrelated to the severity of the anoxic brain injury. The elimination of scans deemed to be unfit for quantitative analysis was based on visual assessment, rather than strict a prior criterion. In particular, the existence of significant white matter disease introduces selection variability, as this is a common finding of varying severity in this population. However, steps were taken to ensure that assessment of scans were performed blinded to the patient’s clinical outcome.

## Conclusion

In summary, quantitative methodologies offers an objective and replicable evaluation of the severity of anoxic brain injury after cardiac arrest. At present, their accuracy is inferior to radiological interpretations, even when considering the potential bias of the self-fulfilling prophecy. Our research indicates that a significant portion of this discrepancy arises from issues like scanner artifacts, chronic strokes, patient motion, and white matter pathology. Future investigations should focus on refining these quantitative methods, especially in terms of adjusting for pre-existing conditions and ensuring optimal scan quality.

## Data Availability

All data produced in the present study are available upon reasonable request to the authors

## Notes

Conflicts of Interest: Jong Woo Lee: co-founder (Soterya Inc); contract work (Teladoc); consultant (SK Biopharm); research funding (NIH/NINDS; UCB/Parexel)

### Competing Interest Statement

The authors have declared no competing interest.

### Funding Statement

This study did not receive any funding

